# Artificial intelligence-assisted ganglion cell detection in Hirschsprung’s disease: A comparative evaluation of two deep learning approaches

**DOI:** 10.64898/2026.06.11.26354826

**Authors:** Eve Wang, Karl Grenier, Peter Savadjiev, Dan Poenaru

## Abstract

**Background:** Definitive diagnosis of Hirschsprung’s disease (HD) requires pathological identification of enteric ganglion cells. This process is time-consuming and subject to inter-observer variability. Artificial intelligence (AI) tools have the potential to standardize and accelerate this workflow, but no study has determined which AI approach best serves intraoperative HD pathology diagnostics.

**Method:** This study compared the U-Net and You Only Look Once version 26 (YOLO26) frameworks for ganglion cell detection using a single-centre retrospective dataset of 54 whole-slide images (WSIs) from rectal biopsies. WSIs were tiled into 397,731 image patches (128×128 pixels), further partitioned into training (70%), validation (15%), and testing (15%) sets. Models were evaluated on tile- and patient-level diagnostic metrics and processing latency.

**Results:** The U-Net achieved a tile-level sensitivity of 82.9%, showing no statistically significant difference compared to YOLO26 (79.1%; p = 0.097). However, YOLO26 demonstrated a statistically significant advantage in tile-level specificity (96.1% vs. 93.9%; p < 0.001) and reduced mean inference latency (7.64 ms vs. 11.57 ms/tile). At the patient level, both models achieved 100% diagnostic sensitivity. Despite low patient-level specificity (0.0% U-Net; 11.8% YOLO26), the tissue-level diagnostic burden of false positives was 6.00% for U-Net and 3.50% for YOLO26.

**Conclusion:** The U-Net is preferred when nominal gains in sensitivity are prioritized, while the YOLO26 is an alternative that optimizes efficiency and false positive suppression. Both models serve as robust screening filters to augment the pathologist’s workflow and should be selected based on workflow requirements. Prospective validation on larger, multi-centre datasets is required before clinical implementation.

## 1. Introduction

Hirschsprung’s disease (HD) is a rare congenital disorder characterized by the absence of ganglion cells in the distal colon. Occurring in approximately 1 in 5,000 live births, the condition results from the failure of neural crest cells to migrate completely during embryonic development (1). This absence leads to chronic obstruction and is potentially fatal if left untreated (2). Consequently, rapid and accurate diagnosis is critical for timely surgical intervention and patient survival.

The current gold standard for diagnosing HD involves the histopathological examination of rectal biopsy specimens (3). The diagnostic process ultimately becomes a laborious search, in which the pathologist must identify even one ganglion cell to rule out the disease. However, identifying ganglion cells can be difficult without specific training. While artificial intelligence (AI) offers a potential solution, the optimal architectural paradigm for this task remains an open research question.

Recent advancements in deep learning and whole-slide images (WSIs) have enabled the growth of computational pathology applications. By digitizing histology slides, computer vision algorithms can consistently analyze tissue samples, potentially reducing the risk of false negatives. Currently, semantic segmentation architectures (e.g., U-Net (4)) are commonly used in HD computational pathology (5, 6). These architectures classify every pixel in an image, provisioning precise morphological boundaries of cells. While this offers granular detail, it is computationally expensive and requires rigorous pixel-level annotations (7). Conversely, object detection architectures such as You Only Look Once (YOLO) (8) generate bounding boxes rather than pixel maps and are rarely applied in computational pathology. These models are significantly faster and require less granular annotations.

This paper presents a comparative analysis of these two approaches configured for an intended clinical role as a high-sensitivity screening and triage tool to assist pathologists in ruling out aganglionosis. By benchmarking a U-Net-based segmentation model against a YOLO-based detection model, we aim to evaluate the trade-offs between diagnostic sensitivity, specificity, and computational efficiency. We hypothesized that while both deep learning frameworks would achieve comparable tile-level sensitivity, the object detection paradigm would yield a significant reduction in processing latency, thereby optimizing pathologist review workflows without compromising diagnostic safety. Our goal is to determine whether rigorous pixel-wise segmentation analysis is required for clinical accuracy or if rapid object detection inference is sufficient for effective screening.

## 2. Methods

The goal of this exploratory, comparative study was the creation and validation of deep learning pipelines intended for a clinical role in high-sensitivity screening. Specifically, predictive modeling targeted the localized detection of ganglion cells to accelerate the ruling out of aganglionosis in tissue sections. The development and performance evaluation of our deep learning models were reported in accordance with the Checklist for Artificial Intelligence in Medical Imaging (CLAIM) 2024 guidelines (9, 10).

### 2.1 Dataset Preparation

This retrospective cohort study utilized a dataset consisting of 54 previously digitized HPS-stained pathology slides of all rectal biopsies performed for the diagnosis of HD in children 0-10 years at the Montreal Children’s Hospital between 2015 and 2022. All WSIs were anonymized by removing patient identifiers before downstream processing. Ganglion cells were annotated using QuPath (11) by a medical student for a primary screening, and the annotations were subject to a secondary verification by a senior pediatric pathologist.

To handle the large sizes of the WSIs, the slides were tiled into 397,731 128×128 pixel tiles with 50% overlap. Tiles that were of low quality (i.e., 50%+ white space, blurry, and contained slivers of ganglion cell annotations) were removed manually. The tiles were subsequently split into training (70%), validation (15%), and testing (15%) sets, ensuring patient-level splits to prevent data leakage. Due to class imbalance in the training set, the majority class was downsampled, and the minority class was oversampled to achieve a 30:70 ratio of background to ganglion cell images. Data augmentation techniques, including image rotation, flipping, and colour jittering, were applied to the training set to improve model generalization. Different stain normalizations were used to align with the different architectures and preserve pre-trained weight integrity. The U-Net input was normalized using Z-score normalization to match the expected input distribution of its ImageNet-pretrained encoder weights (12). Conversely, the YOLO26 model utilized its native Min-Max scaling ([0,1]), which is handled internally by the Ultralytics inference engine to align with its COCO-pretrained baseline. COCO, short for Common Objects in Context, is a large-scale dataset containing images of common objects, often used for segmentation and computer vision tasks (7). Deviating from these native pipelines would introduce severe data distribution shifts, degrading the feature-extraction capabilities of the respective pre-trained backbones.

### 2.2 Model Architectures

Two distinct model architectures were implemented: a U-Net (4) for semantic segmentation and a YOLO26 (You Only Look Once version 26) (13) model for object detection (**Fig. 1**).

**Fig. 1.**
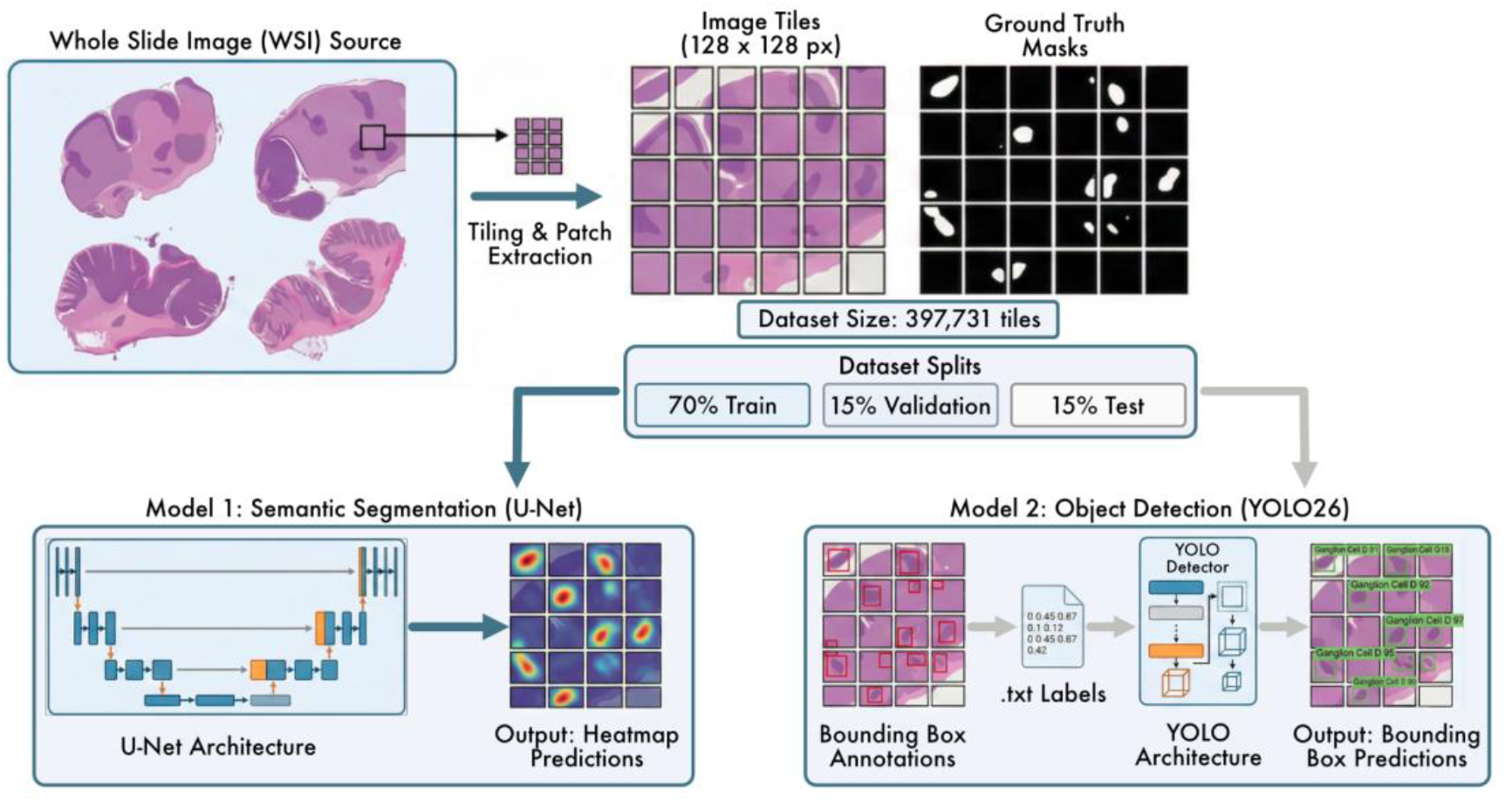
Overview of the comparative methodology for the computer-aided pathological diagnosis of Hirschsprung’s disease. Whole-slide images (WSIs) were pre-processed by tiling into overlapping 128×128 pixel patches, yielding a total dataset of 397,731 tiles. The tiles were split into training (70%), validation (15%), and testing (15%) sets. The study evaluated two architectures: Model 1 (Left) employed a U-Net architecture for semantic segmentation and generated pixel-wise probability heatmaps of ganglion cells. Model 2 (Right) employed a YOLO26 architecture for object detection, using bounding-box annotations to localize cells. Both models were evaluated on the same test set to compare diagnostic sensitivity and inference efficiency.

The U-Net architecture was selected for its successful application in biomedical image segmentation. It uses symmetric encoder-decoder blocks that create its distinct “U” shape. For feature extraction, we used an EfficientNet-B3 (14) backbone as the encoder, initialized with ImageNet pre-trained weights (12). This backbone leverages compound scaling and squeeze-and-excitation blocks to capture complex morphological features at multiple scales. The decoder uses up-convolutions to restore spatial resolution. Skip connections pass high-resolution features from the encoder directly to the decoder for feature preservation. The final layer uses a 1×1 convolution with a sigmoid activation to generate the segmentation mask.

For object detection, the YOLO26 (medium) was selected for its optimized deployment efficiency and end-to-end performance. The model uses a CSP-Darknet-style (15) backbone for feature extraction and a Path Aggregation Network (PANet) (16) neck for multi-scale feature fusion. Unlike previous iterations, YOLO26 employs a native end-to-end predictor that eliminates the need for Non-Maximum Suppression (NMS), significantly reducing inference latency while maintaining high sensitivity. This model was initialized with COCO (7) pre-trained weights and fine-tuned on our patient-derived dataset.

### 2.3 Training and Implementation

The U-Net was implemented in Python using the segmentation_models.pytorch library (17), and the YOLO26 model was implemented using the Ultralytics library (13). Training was conducted on an NVIDIA RTX 5000 GPU. Model hyperparameters were optimized using a randomized grid search, with the final selected values detailed below.

The U-Net was optimized using AdamW with a scheduler, an initial learning rate of 1e-4, and a batch size of 32. The model was trained for 100 epochs. The loss function was defined as a weighted combination of Dice Loss and Focal Loss, with 30% of the weight assigned to Dice Loss and 70% to Focal Loss, and a 0.5 alpha value for the Focal Loss. The best model was selected based on the lowest validation loss during training.

The YOLO26 was trained for 100 epochs using the AdamW optimizer with a learning rate scheduler, starting with an initial rate of 1e-4 and a batch size of 32. Optimization was driven by its native training engine configured without ensembling techniques. The loss was minimized using the standard YOLO26 multi-part loss function, and the best model was selected based on a fitness metric where:

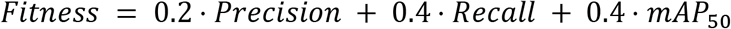

Where mAP_50_ was a metric calculated automatically by the Ultralytics library. This metric determined whether the overlapping bounding box covered at least 50% of the area of the ground truth box.

### 2.4 Model Performance Evaluation

Probability heatmaps for the U-Net and bounding boxes for the YOLO26 served as the primary mechanism for model interpretability, enabling qualitative evaluation of model outputs. To quantitatively evaluate the performance of both models, we implemented a custom probing method for tile-level predictions. Rather than relying on pixel-wise metrics, this metric translates model outputs into a detection problem.

#### 2.4.1 Model Standardization

For a direct comparison between the U-Net and YOLO26, all outputs were standardized to a bounding-box format before evaluation, with values selected based on a sensitivity analysis.

For the U-Net framework, predicted continuous probability maps were binarized. Connected component labeling was subsequently applied to the resulting binary masks to isolate distinct regions, and the spatial extremities of each discrete pixel cluster were extracted to construct the associated bounding box coordinates. Conversely, the YOLO26 model’s native bounding box coordinate outputs were used directly.

#### 2.4.2 The Probe Algorithm

The custom algorithm subjected the standardized bounding boxes to a rigorous three-point structural validation process to determine each tile’s diagnostic status relative to the ground truth. To ensure these threshold selections did not systematically bias evaluation toward either architecture, a sensitivity analysis was performed across a spectrum of parameter spaces. The resulting optimization data are provided in Supplementary Table 1 and Supplementary Table 2.

1. Size Match: For the U-Net, an inside signal ratio was computed, requiring the binarized pixel prediction to span at least 20% of the ground truth target area. For YOLO26, this logic was symmetrically mirrored using an Intersection over Union (IoU) probe, requiring a spatial overlap threshold of > 0.2 to register a successful detection.
2. Alignment: The Euclidean distance between the predicted center of mass and the ground truth centroid was measured. A relative offset >15% of the box diagonal resulted in an off-center penalty.
3. Noise Discrimination: To prevent the artificial inflation of accuracy metrics from over-segmentation, an anti-cheat check measured external noise levels for the U-Net. If more than 15% of the target box’s immediate surrounding peripheral area contained predicted positive pixels, the detection was flagged as a false positive.

#### 2.4.3 Diagnostic Status Classification

Based on the results of the structural probe, each tile was assigned to one of the following: True Positive (TP), where the prediction passed all checks; False Positive (FP), where the model hallucinated a structure or failed the noise discrimination check; False Negative (FN) if the ground truth structure was present but the model failed to generate a detection or failed the size match test; True Negative (TN) if the ground truth and model correctly predicted an empty tissue region.

#### 2.4.4 Evaluation Metrics

Following the classification of tiles into TP, FP, FN, and TN, we calculated four primary statistical metrics to evaluate diagnostic performance: sensitivity, specificity, balanced accuracy, and inference time per tile in ms. Balanced accuracy was used as an alternative to standard accuracy, given the potential for class imbalance between positive and negative tiles, and was calculated as follows:

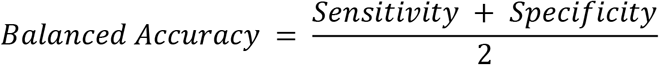

To statistically evaluate differences in model performance at the tile-level, paired McNemar’s tests were applied independently to sensitivity and specificity by stratifying the dataset based on ground truth. To isolate sensitivity, the test was restricted exclusively to ground truth positive cases, evaluating the discordance between the TPs and FNs on an image-by-image basis. Analogously, to isolate specificity, a separate McNemar’s test was performed on ground truth negative cases to compare the model’s discordant TN and FP classifications. For both, a two-tailed p-value of <0.05 was used to denote statistical significance.

Patient-level metrics were measured based on patch-level aggregations. The number of predicted ganglion cells for each patient was recorded and compared against a ground truth diagnosis. 95% confidence intervals (CI) were calculated using the Wilson Score method.

### 2.5 Environmental Footprint

To assess the environmental impact of model training and inference, we quantified the carbon footprint and energy consumption for both the U-Net and YOLO26 architectures using the AI Environmental Footprint Calculator (18). The calculation was based on various hardware inputs and model parameters. To ensure regional accuracy, the carbon intensity was mapped to the Montreal, Quebec, Canada, power grid.

## 3. Results

### 3.1 Quantitative Performance

The study comprised a total of 54 pediatric patients distributed across disjoint data splits. Patient-level partitioning allocated 23 patients to the training set, 9 patients to the validation set, and 22 independent cases to the final test pool. The models were evaluated using the tile-level probing framework and patient-level aggregation logic previously defined. Both models were tested on the same held-out test set of 61,184 tiles. Performance metrics for the two architectures are summarized in **Table 1**.

**Table 1.**
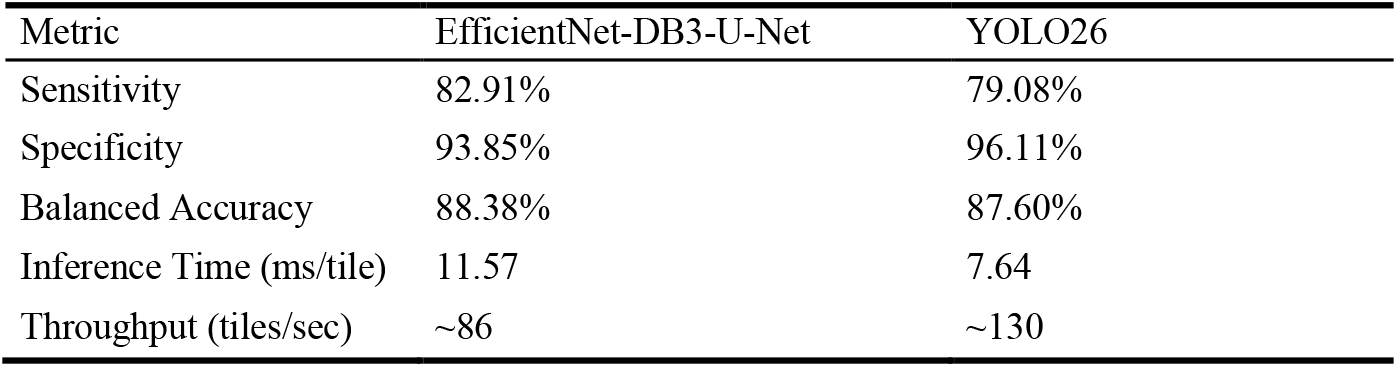
Summary of the quantitative performance metrics for the U-Net and YOLO26.

#### 3.1.1 Tile-Level Performance

Model performance at the tile level was quantified using confusion matrices (**Fig. 2**), from which sensitivity, specificity, and balanced accuracy measures were derived. The U-Net architecture achieved a diagnostic sensitivity of 82.9%, whereas the YOLO26 model demonstrated a slightly lower result at 79.1%. Statistical analysis indicated that the difference in sensitivity between the two models was not statistically significant (p = 0.097), establishing comparable diagnostic performance in ganglion cell localization.

**Fig. 2.**
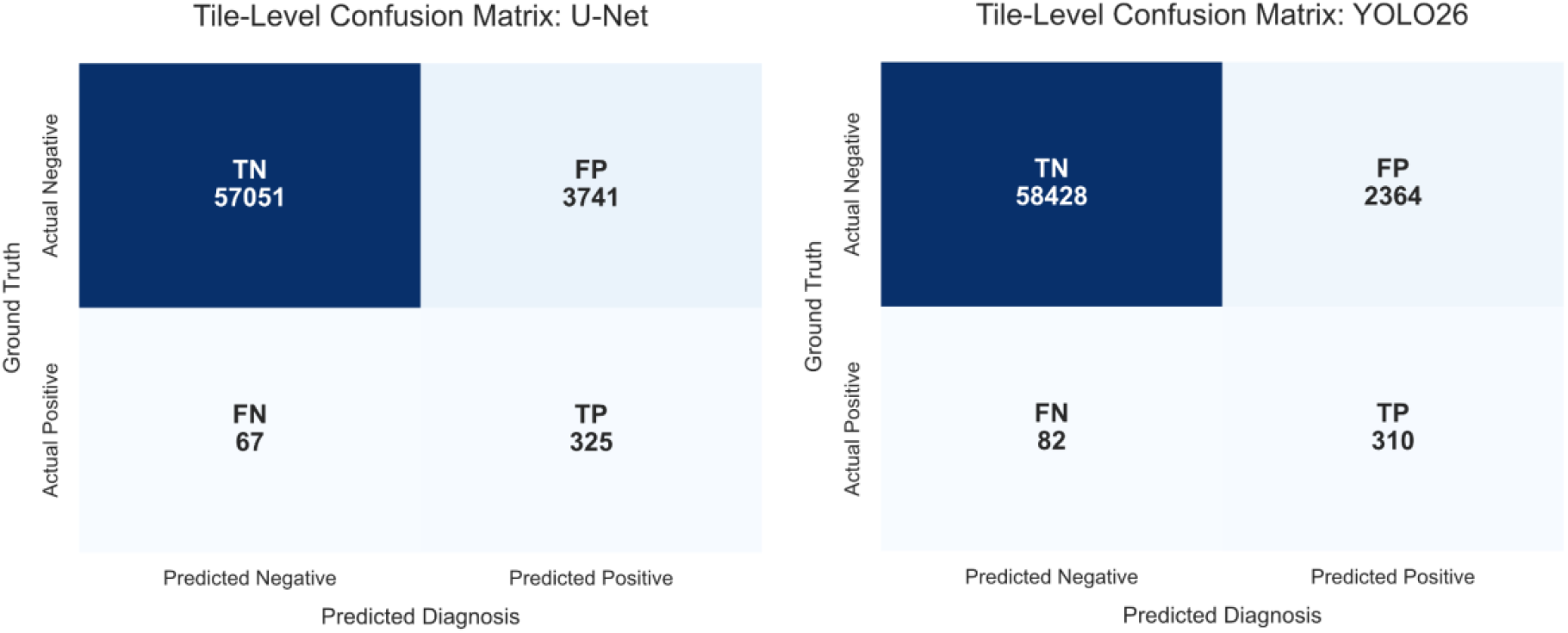
Tile-level confusion matrices for the U-Net and YOLO26 frameworks. TN: True Negative, FP: False Positive, FN: False Negative, TP: True Positive.

Conversely, a statistically significant variation was observed regarding model specificity. The YOLO26 model demonstrated a distinct advantage, achieving a specificity of 96.1% compared to 93.9% for the U-Net (p < 0.001). This variance corresponds to a 36% relative reduction in the false positive rate for the YOLO26 architecture compared to the U-Net. Due to the inverse distribution of these metric advantages, the overall balanced accuracy remained highly comparable between the two architectures.

Computational efficiency was benchmarked by evaluating the mean inference latency required to process an individual 128×128 tile. The YOLO26 demonstrates a mean inference latency of 7.64 ms per tile. In comparison, the U-Net architecture exhibited a mean inference latency of 11.57 ms per tile. This represents a 33.97% reduction in processing time per computational unit in favour of the YOLO26 model.

#### 3.1.2 Patient-Level Performance

To evaluate clinical utility, model predictions were aggregated from individual tiles to the patient level (n = 22). Both the U-Net and YOLO26 architectures achieved 100% patient-level sensitivity (95% CI: 56.6% - 100.0%), successfully identifying all histologically confirmed positive cases (n = 5) with no FNs (**Fig. 3**).

**Fig. 3.**
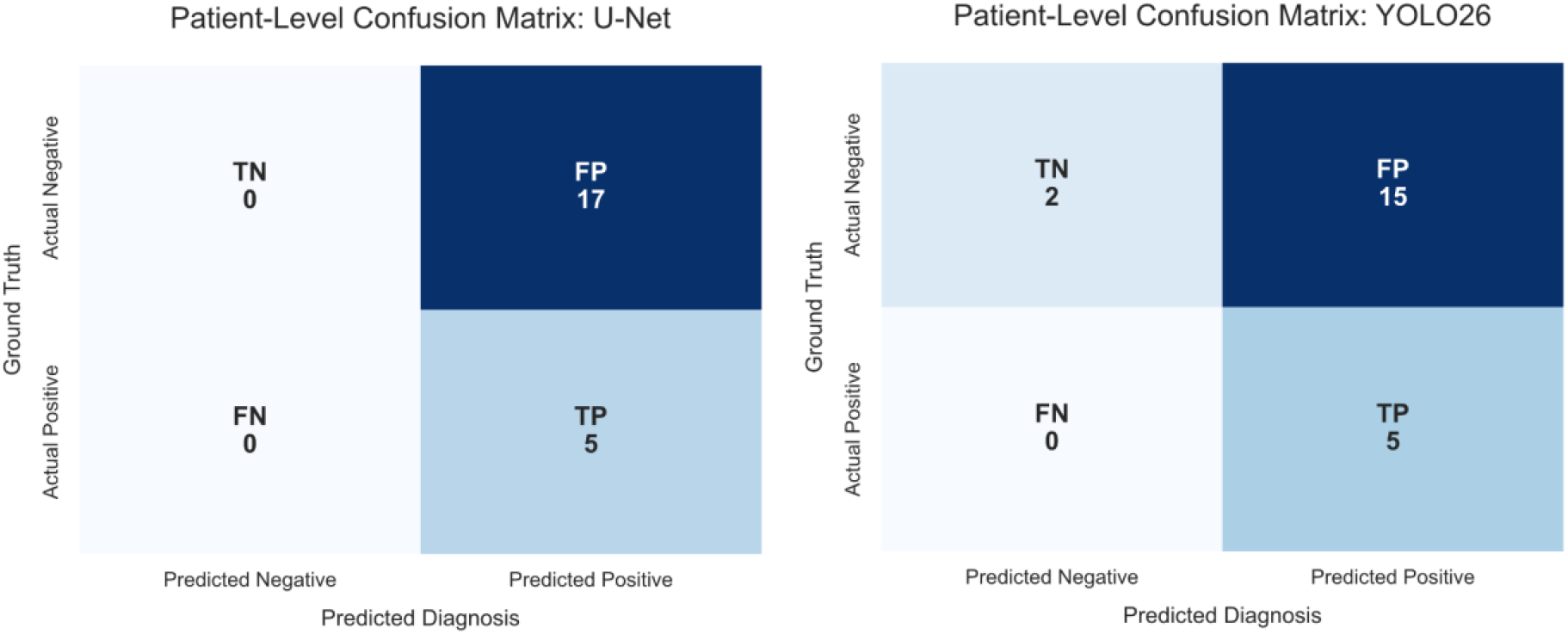
Patient-level confusion matrices for the U-Net and YOLO26 frameworks. TN: True Negative, FP: False Positive, FN: False Negative, TP: True Positive.

When evaluated at a strict single-tile detection threshold, where a single positive prediction flags an entire case, patient-level specificity was low across both frameworks. The U-Net model correctly identified 0 out of 17 negative cases (0% specificity; 95% CI: 0.0% - 18.4%), identifying at least one FP tile in every negative biopsy. The YOLO26 model correctly classified 2 out of 17 negative cases, yielding a patient-level specificity of 11.7% (95% CI: 3.3% - 34.3%).

An evaluation of the total diagnostic burden across the entire histologically negative cohort in the test set (n = 17; total tiles = 37,919) revealed that these FP classifications were sparse. For the U-Net, a total of 2,274 tiles were misclassified as FPs, representing an overall tissue-level FP proportion of 6.00% and an average of 133.8 targeted tiles per patient slide for manual review. For the YOLO26, the total FP count was reduced to 1,328 tiles, corresponding to an overall tissue-level proportion of 3.50% and an average of 78.1 tiles per patient slide. Within a clinical workflow, this distribution requires a pathologist to manually audit only a minute fraction of the total tissue area to confirm a diagnosis.

### 3.2 Qualitative Analysis and Error Characterization

The high sensitivity of both models was further investigated through a qualitative review of the detection outputs. While both models identified the primary diagnostic targets, they exhibited distinct, recurring failure modes when met with complex tissue environments or technical artifacts.

#### 3.2.1 Morphological Successes and Identification

Figure 4. illustrates the predictive performance of both architectures across a spectrum of histological complexities. **Figure 4A** highlights characteristic detection cases, where the ganglion cells exhibit prominent cytoplasmic volume and are predominantly located within the image tile. **Figure 4B** presents more challenging diagnostic scenarios, characterized by smaller cell diameters and peripheral positioning that results in the truncation of the cell boundary. In cases containing ganglion cells (Rows 1-3), both models appear to successfully locate the target cell, with the U-Net producing a pixel-precise segmentation mask and the YOLO26 generating a bounding box centered on the cell body. Furthermore, both models correctly identified the absence of ganglion cells in the negative control (Row 4).

#### 3.2.2 Analysis of False Positives

To characterize the models’ most significant diagnostic failures, an experienced pediatric pathologist conducted a qualitative review of 20 select examples from the FP pool (**Table 2**). These cases were instances where the models assigned a high probability to a detection despite the absence of a ground truth annotation. Understanding these high-confidence errors is vital for clinical safety, as they represent the most likely source of overdiagnosis. In evaluating these errors, a distinction must be made between technological artifacts and true biological mimics. While FPs occurring near edges or were blurry often limit analysis of post-hoc speculation regarding feature extraction errors, misclassifications on structures like smooth muscle, mucosal crypts, or blood vessels represent concrete biological mimics where the tissue shares morphological similarities with the ganglion cell. A detailed breakdown of these specific failure modes for each architecture is provided below.

**Fig. 4.**
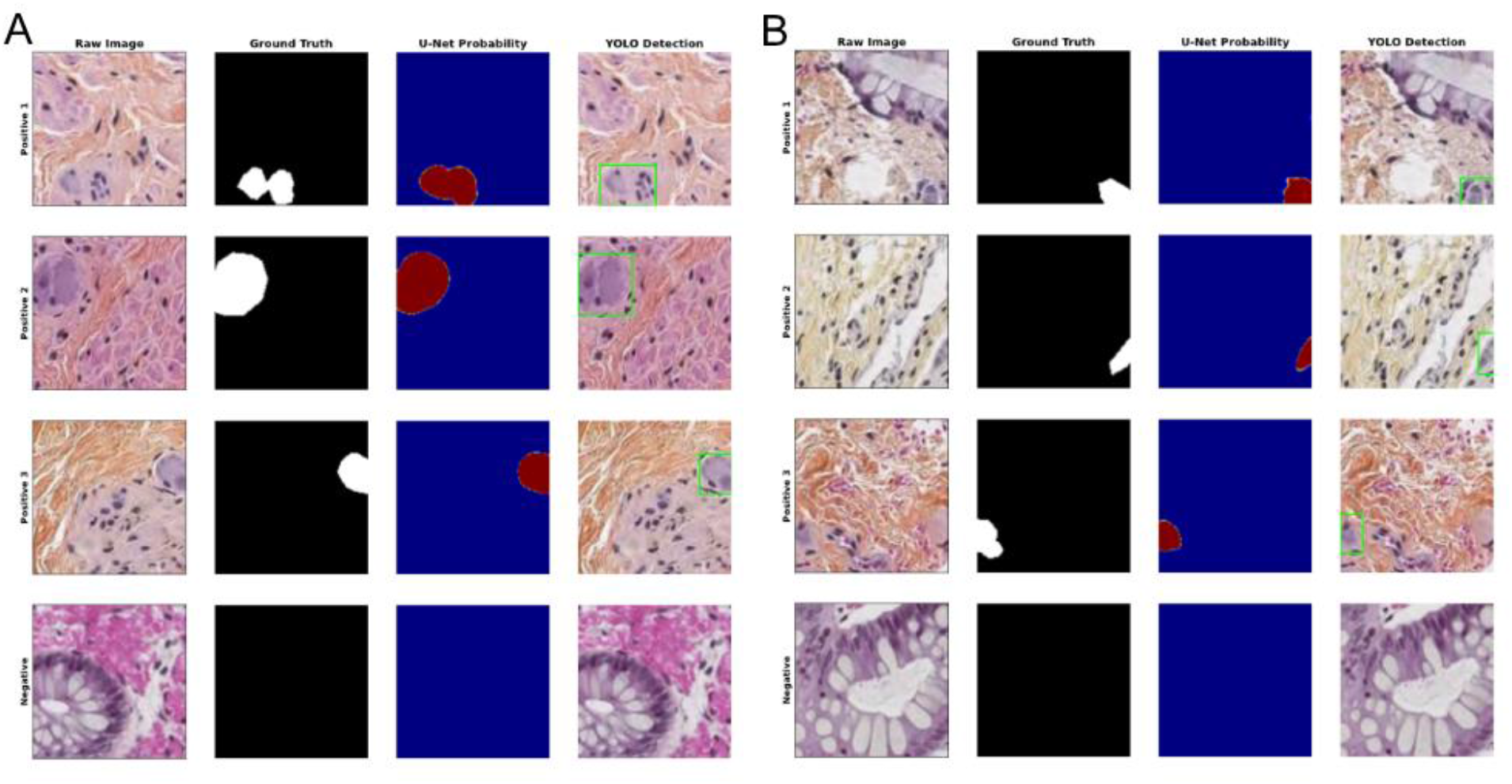
Visualizations of the model predictions. From left to right: raw image, ground truth binary mask, U-Net probability heatmap prediction, YOLO26 bounding-box prediction. **A**. Visualizations of larger ganglion cells. **B**. Visualizations of more difficult edge cases where cells are cut off and smaller in size.

**Table 2.** Summary of the False Positive Morphological Categories of 20 Examples.

| Morphological Category | EfficientNet-B3-U-Net | YOLO26 |
| --- | --- | --- |
| Muscle Tissue | 6 | 12 |
| Possible Ganglion Cell Clusters | 4 | 3 |
| Image Artifacts | 4 | 1 |
| Other | 6 | 4 |

Elaborations: Muscle Tissue (e.g., smooth, vessel), Image Artifacts (e.g., blurry, edge), Other (glands, mast, inflammatory cells)

The U-Net model’s high-confidence FPs were distributed across several morphological categories. Four instances were classified as possible ganglion cell clusters; these regions contained cells with morphological features consistent with neuronal cytoplasm but lacked corresponding manual annotations in the ground truth. Another four instances were associated with image artifacts, specifically localized blurring or unclear tissue edges. The remaining errors were distributed between mucosa crypts (n = 3) and various forms of smooth muscle (n = 6), with the remaining three detections occurring in miscellaneous non-neuronal structures.

The YOLO26 model’s high-confidence error profile was dominated by the misclassification of stromal elements. Over half of the analyzed examples (n = 12) were localized to smooth muscle or vessel smooth muscle. Unlike the U-Net, YOLO26 identified fewer FPs in areas of image blurring (n = 1) but demonstrated a higher frequency of detections in small-cell populations, specifically mast cells and inflammatory cell clusters (n = 4). Three instances were categorized as possible ganglion cell clusters, and the final four errors occurred in miscellaneous vascular and glandular structures.

#### 3.2.3 Analysis of False Negatives

A comparative analysis was conducted to identify recurring patterns in FN detections, which were instances where the models failed to identify specific ganglion cells present in the ground truth annotations. A total of 67 and 82 FNs were identified for the U-Net and YOLO26 models, respectively. Each omission was reviewed and categorized based on local image quality and surrounding histological context.

The primary source of error for both architectures was related to image quality, with many omitted cells exhibiting multiple overlapping conditions (e.g., being both blurry and located at the edge of a tile). Blurry regions accounted for the largest share of FNs (n = 39 for U-Net; n = 37 for YOLO26), followed by cells located at the edge of the 128×128 pixel tiles (n = 20 for U-Net; n = 21 for YOLO26). Suboptimal lighting conditions, categorized as dark regions, contributed to 10 U-Net and 13 YOLO26 omissions. Beyond technical artifacts, proximity to smooth muscle was associated with 10 U-Net FNs and 3 YOLO26 FNs, while proximity to white areas (representing clear space or lipid content) was associated with 7 and 4 FNs, respectively.

Notably, every ground truth ganglion cell missed by the models occurred in cases where other ganglion cells within the same biopsy had been correctly identified. Consequently, while these instances were recorded as FNs on a cell-by-cell basis, the models achieved a 100% correct diagnosis at the patient level. In every case reviewed, the presence of at least one detected ganglion cell would have resulted in a clinical diagnosis of “negative for HD,” matching the pathologist’s final assessment.

### 3.3 Environmental Impact

To evaluate computational sustainability, the environmental impact of model training and clinical inference was assessed (detailed in Supplementary Table 3 and Supplementary Table 4). While the U-Net required less energy to train (1.60 kWh vs. 2.94 kWh), the YOLO26 framework was more efficient during inference (0.058 kWh vs. 0.088 kWh), highlighting its potential for scalable, energy-efficient clinical deployment.

## 4 Discussion

The diagnosis of HD remains a significant challenge due to the labor-intensive nature of identifying the absence of ganglion cells (19). This study establishes a baseline for assisted diagnosis by comparing pixel-wise semantic segmentation (U-Net) against single-stage object detection (YOLO26). Unlike typical histopathological diagnoses, HD is a diagnosis of exclusion that requires confirmation of *absence* rather than *presence*. Computer vision tools are ideal for this task because they can maintain constant scrutiny over larger datasets; however, they must be highly sensitive to ensure that ganglion cells are not missed.

The broader digital pathology literature demonstrates that AI integration is heavily dictated by clinical context. In rare pediatric diseases, extreme data scarcity forces architectures to optimize for accuracy (20), whereas in intraoperative environments, clinical utility is driven largely by operational turnaround time and seamless workflow integration (21). Existing applications by Duci et al. and Oda et al. have demonstrated the U-Net’s effectiveness for HD diagnosis, with Duci et al. (5) reporting an accuracy of 92.3% and Oda et al. (6) reporting a sensitivity of 85.3%. While our U-Net achieved comparable performance, our work highlights a critical oversight in the literature: the trade-off of annotation costs and inference latency. Because maintaining high sensitivity is the non-negotiable priority for HD screening, the question shifts to which architectural framework optimizes the pathologist’s auditing time. By evaluating these processing constraints alongside clinical accuracy, we demonstrate that the model’s value lies in its capacity to accelerate the human-AI workflow.

While Rong et al. (22) evaluated the YOLO architecture for the distinct task of nucleus segmentation, our findings indicate that the model’s performance characteristics remain consistent when applied to ganglion cell detection in HD. Specifically, our observed recall metrics align with their results established in their oncological study, suggesting that YOLO26’s feature-extraction capabilities are robust across varying tissues. When comparing the YOLO26 and U-Net models, we found that YOLO26 demonstrated a 34% reduction in inference latency and a 41.6% absolute reduction in the total FP tile burden (1,328 tiles vs. 2,274 tiles). This performance translated to a lower tissue-level FP proportion (3.50% vs. 6.00%) and fewer average flagged slides (78.1 vs. 133.8). This suggests that, while demonstrating statistically comparable sensitivity to U-Net, YOLO26’s speed and false positive suppression make it a viable candidate for rapid clinical triage, provided that a pathologist reviews the prediction. Regarding sustainability, the energy consumption for both models remained relatively low during inference. Although the YOLO26 model has more parameters and requires more training energy, the inference energy per image is lower than that of the U-Net. Ultimately, model selection should be dictated by specific clinical priorities. The U-Net provides a slightly higher sensitivity that may make it preferable in critical surgical windows, despite its higher manual review workload. Conversely, the YOLO26 offers a highly scalable alternative for routine, high-volume workflows where deployment energy savings and significant FP suppression are prioritized.

While direct diagnostic accuracy comparisons between AI and pathologists are common, the more critical metric for clinical integration should be the cooperation between the two. Based on our findings, the models achieved a notable reduction in per-tile latency, suggesting that AI-assisted review can significantly decrease the time required for a pathologist to review a slide. Considering that the manual screening of a rectal biopsy WSI without ganglion cells takes three to five minutes for our pathologist (KG), the introduction of either AI model could reduce this to a fraction of the time by highlighting only the high-probability candidates for review. These models would be most valuable in true HD cases, where cells are critically sparse, and pathologists spend the most time. By flagging a targeted subset of candidate regions, including both TPs and FPs, this allows the pathologist to focus strictly on the ambiguous area rather than having to scan the entire tissue. In our dataset, the median slide density was approximately 4,000 tiles per patient. At this scale, the total raw computational inference time required to process an entire patient case spans just 30.56 seconds for the YOLO26 framework (4,000 tiles x 7.64 ms) and 46.28 seconds for the U-Net architecture (4,000 tiles x 11.57 ms). This rapid processing shifts the clinical workflow from an exhaustive, manual pixel-by-pixel search across thousands of fields of view into a highly directed auditing task completed in under a minute, drastically shrinking the diagnostic timeline.

However, this transition from searching to auditing necessitates a cautious approach. Given that both architectures still produced hallucinated artifacts, the pathologist’s review remains the indispensable ground truth. This human-in-the-loop requirement ensures that the AI functions as a safety multiplier rather than an autonomous diagnostic agent. The importance of this collaborative dynamic is underscored by our error analysis, which revealed that several false positives produced by the U-Net were, upon secondary review, suspicious cellular clusters with morphological features suggestive of ganglion cells. In these instances, the model’s pixel-level sensitivity allowed it to flag potential targets that fell outside the initial, more conservative manual annotations. Moreover, the tool can help identify ganglion cells in the transition zone, where there can be partial circumferential aganglionosis and myenteric hypoganglionosis. This suggests that the framework does not merely replicate human effort but may act as a high-sensitivity fail-safe, surfacing subtle biological signals that warrant closer clinical inspection (23). Furthermore, the pathologist provides a layer of global contextual synthesis that the models currently lack, such as correlating the presence of ganglion cells with the patient’s age, clinical history, and biopsy location (24). The value of the model should thus be measured by its ability to mitigate observer fatigue and standardize the screening process. By automating the most tedious aspects of the search, this framework allows the pathologist to focus their expertise on the most complex borderline cells, thereby enhancing diagnostic rigor and safety of the HD diagnostic workflow.

Several limitations likely constrained the performance of our models. Our dataset prioritized scale over curation, introducing label noise through low-quality “edge case” tiles. Additionally, because this study was executed as an exploratory evaluation of internal data splits, the models were not validated against an external multi-centre dataset, nor was the study registered as a prospective clinical trial. Converting U-Net maps to bounding boxes for comparison also reduced rich morphological data to coarse coordinates, potentially penalizing U-Net’s latency while failing to capture its diagnostic value. Finally, relying on tile-level metrics creates a bottleneck for clinical integration, as high tile accuracy does not guarantee success in patient-level diagnostics (25). High sensitivity remains vital to ensure that, in borderline cases where ganglion cells are exceptionally sparse, the model can successfully identify the few cells that are present. Qualitatively, both models are disadvantaged by a lack of global contextual information. Pathologists often rely on secondary clues, such as the presence of hypertrophic submucosal nerves, to increase diagnostic certainty, which is context that is currently lost in localized 128×128 tile-level processing (26).

Future work will focus on prospective validation on larger, multi-centre datasets to ensure the models remain robust across diverse laboratory protocols and digital scanning platforms. Crucially, this must include the evaluation of intraoperative frozen-section material, where the models’ sensitivity must be tested against unique histological challenges. To quantify the clinical utility of these tools, further studies should benchmark performance across three different cohorts: the model operating autonomously, the model assisting specialist pediatric pathologists, and the model supporting general pathologists. This comparison would help determine the degree of diagnostic security provided and the time saved in the workflow. Additionally, future work will investigate the use of global contextual information, such as incorporating the presence of hypertrophic submucosal nerves into the decision-making logic or processing WSIs. The focus will transition toward optimizing performance for patient-level diagnosis rather than individual tiles, ensuring that the AI’s output aligns directly with the binary diagnostic requirements of clinical pathology.

## 5. Conclusion

This study compared semantic segmentation (U-Net) and object detection (YOLO26) for ganglion cell detection in HD. While both models demonstrated statistically comparable diagnostic sensitivity, their clinical utility is defined by a distinct trade-off between architectural precision, FP suppression, and operational efficiency. The U-Net provides a high sensitivity, suitable for critical surgical windows, whereas the YOLO26 framework offers a high-speed, deployment-efficient alternative, making it viable for large-scale clinical screening. Importantly, error analysis revealed that every cell missed by either model occurred in a biopsy where other ganglion cells were successfully identified, yielding 100% patient-level sensitivity across both models. By establishing that the diagnostic process can be successfully reframed as an AI-assisted filtering task, this study provides a flexible framework for modern digital pathology. Ultimately, the integration of these models serves to augment the pathologist’s workflow rather than replace it, mitigating the risks of observer fatigue and standardizing diagnostic rigor to ensure a higher margin of safety and efficiency in the diagnosis of HD.

## Data Availability

The source code developed in this study is publicly available in the GitHub repository. The de-identified clinical datasets and histopathological images analyzed during the current study are not publicly available due to institutional data privacy regulations and patient confidentiality agreements.

https://github.com/yixuanevewang/Hirschsprungs-disease-model-comparison

## Funding

This research received no specific grant from any funding agency in the public, commercial, or not-for-profit sectors.

## Competing Interest Statement

The authors have no conflicts of interest to declare.

## Ethical Approval

This study was approved by the Research Ethics Board of McGill University Hospital Centre (MUHC).

## Notes

### Competing Interest Statement

The authors have declared no competing interest.

### Author Declarations

Research Ethics Board of McGill University Health Centre gave ethical approval for this work.

