## Supplementary Table 1 for "Artificial intelligence-assisted ganglion cell detection in Hirschsprung’s disease: A comparative evaluation of two deep learning approaches"

**Supplementary Table 1.** U-Net Sensitivity Sweep for Probe Algorithm

| Probability Threshold | Min Signal Ratio | Max Noise Ratio | Max Offset | Centre | Sensitivity |
|---|---|---|---|---|---|
| 0.2 | 0.2 | 0.05 | 0.15 | | 0.853 |
| 0.2 | 0.2 | 0.05 | 0.25 | | 0.853 |
| 0.2 | 0.2 | 0.05 | 0.35 | | 0.853 |
| 0.2 | 0.2 | 0.1 | 0.15 | | 0.855 |
| … | … | … | … | | … |
| **0.2** | **0.2** | **0.15** | **0.15** | | **0.857** |
| … | … | … | … | | … |
| 0.6 | 0.5 | 0.2 | 0.35 | | 0.761 |

288 configurations were evaluated across various combinations of the four parameters. Selected the combination reporting the highest sensitivity.
