## Supplementary Table 2 for "Artificial intelligence-assisted ganglion cell detection in Hirschsprung’s disease: A comparative evaluation of two deep learning approaches"

**Supplementary Table 2.** YOLO26 Sensitivity Sweep for Probe Algorithm

| Confidence Threshold | Recall Threshold | Sensitivity |
|---|---|---|
| **0.1** | **0.2** | **0.723** |
| 0.2 | 0.2 | 0.688 |
| 0.3 | 0.2 | 0.653 |
| 0.4 | 0.2 | 0.627 |
| 0.5 | 0.2 | 0.610 |
