## Supplementary Table 3 for "Artificial intelligence-assisted ganglion cell detection in Hirschsprung’s disease: A comparative evaluation of two deep learning approaches"

**Supplementary Table 3.** Summary of inputs used to calculate environmental costs for training and inference of both models

| Inputs | EfficientNetB3-U-Net | YOLO26 |
|---|---|---|
| Model Size (Parameters) | 13,159,033 | 21,774,430 |
| Device Utilization (%) | 90 | 90 |
| System Base Power (W) | 95 | 95 |
| Number of Accelerators | 1 | 1 |
| Number of Images (Training) | 6,983 | 6,983 |
| Number of Epochs Trained | 100 | 100 |
| Time/Epoch (Training) (s/epoch) | 128 | 236 |
| Electricity Price ($/kWh) | 0.1 | 0.1 |
| Number of Images (Inference) | 61,184 | 61,184 |
| Time per Image (Inference) (s/image) | 0.01157 | 0.00764 |
