## Supplementary Table 4 for "Artificial intelligence-assisted ganglion cell detection in Hirschsprung’s disease: A comparative evaluation of two deep learning approaches"

**Supplementary Table 4.** Results of environmental impact for the U-Net and YOLO26 architectures during training and inference

| Characteristics | EfficientNetB3-U-Net | YOLO26 |
|---|---|---|
| **Training Phase** | | |
| Energy per image (IT) (kWh) | 1.76e-6 | 3.24e-6 |
| Total IT energy (kWh) | 1.23 | 2.26 |
| Facility energy (IT x PUE) (kWh) | 1.60 | 2.94 |
| Total $CO_2e$ (operational + embodied) (kg) | 0.18 | 0.34 |
| **Inference Phase** | | |
| Energy per image (IT) (kWh) | 1.11e-6 | 7.32e-7 |
| Total IT energy (kWh) | 0.068 | 0.045 |
| Facility energy (IT x PUE) (kWh) | 0.088 | 0.058 |
| Total $CO_2e$ (operational + embodied) (kg) | 0.010 | 0.0067 |
